# Understanding the effects of Universal Test and Treat on longitudinal HIV care continuum outcomes among South African youth: a retrospective cohort study

**DOI:** 10.1101/2022.08.22.22279067

**Authors:** Lindsey M. Filiatreau, Jessie K. Edwards, Nkosinathi Masilela, F. Xavier Gómez-Olivé, Nicole Haberland, Brian W. Pence, Joanna Maselko, Kathryn E. Muessig, Chodziwadziwa Whiteson Kabudula, Mi-Suk Kang Dufour, Sheri A. Lippman, Kathleen Kahn, Audrey Pettifor

**Affiliations:** Department of Psychiatry, School of Medicine, Washington University in St. Louis; International Center for Child Health and Development, Brown School, Washington University in St. Louis; Department of Epidemiology, Gillings School of Global Public Health, University of North Carolina at Chapel Hill, Chapel Hill, North Carolina, USA; MRC/Wits Rural Public Health and Health Transitions Research Unit (Agincourt), School of Public Health, Faculty of Health Sciences, University of the Witwatersrand, South Africa; Population Council, New York, New York, USA; Department of Health Behavior, Gillings School of Global Public Health, University of North Carolina at Chapel Hill, Chapel Hill, North Carolina, USA; Division of Prevention Science, Department of Medicine, University of California, San Francisco, San Francisco, California, USA; Carolina Population Center, Chapel Hill, North Carolina, USA

**Author notes:** Corresponding Author: Lindsey M. Filiatreau, Department of Psychiatry, 660 S. Euclid Ave. St. Louis, MO 63110. E-mail addresses of authors: LMF JKE NM FXG-O NH BWP JM KM CWK MKD SAL KK AP.

**Keywords:** Universal Test and Treat, HIV care continuum, youth living with HIV, South Africa, viral suppression, retention in care

## Abstract

**Introduction:** Little is known about when youth living with HIV (YLHIV) are most susceptible to disengagement from HIV care. The longitudinal HIV care continuum is an underutilized tool that can provide a holistic understanding of population-level HIV care trajectories and be used to compare treatment outcomes across groups. We aimed to explore effects of the Universal Test and Treat policy (UTT) on longitudinal care outcomes among South African youth living with HIV (YLHIV) and identify temporally precise opportunities for re-engaging this priority population.

**Methods:** Using medical record and census data, we conducted a retrospective cohort study among youth aged 18-24 newly diagnosed with HIV between August 2015 and December 2018 in nine health care facilities in rural South Africa. We used weighted Fine and Grey sub-distribution proportional hazards models to characterize longitudinal care continuum outcomes in the population overall and stratified by treatment era of diagnosis. We estimated the proportion of individuals in each stage of the continuum over time and the mean time spent in each stage in the first year following diagnosis. Estimates for the two groups were compared using differences (diagnosis pre-UTT=referent).

**Results:** A total of 420 YLHIV were included. By the end of the first year following diagnosis, just 23% of individuals had no 90-or-more-day lapse in care and were virally suppressed. Those diagnosed in the UTT era spent less time as ART-naïve (mean difference=-19.3 days; 95% CI: - 27.7, -10.9) and more time virally suppressed (mean difference=17.7; 95% CI: 1.0, 34.4) compared to those diagnosed pre-UTT adoption. Most individuals who were diagnosed in the UTT era and experienced a 90-or-more-day lapse in care disengaged between diagnosis and linkage to care or ART initiation and viral suppression.

**Conclusions:** Implementation of UTT yielded modest improvements in time spent on ART and virally suppressed among South African YLHIV. However, meeting UNAIDS’ 95-95-95 targets remains a challenge in this priority population. Retention in care and re-engagement interventions that can be implemented between diagnosis and linkage to care (e.g., longitudinal counseling following diagnosis) or ART initiation and viral suppression may be particularly important to improving treatment outcomes among South African YLHIV.

## Introduction

Traditional methods for monitoring HIV treatment outcomes often fail to capture the complexity of the processes of engagement, disengagement, and reengagement in care that individuals living with HIV may cycle through over time. The longitudinal HIV care continuum framework is an underutilized alternative to these approaches that can provide a more nuanced picture of population-level outcomes over time (1,2). This tool may be particularly useful for assessing holistic differences in treatment outcomes in distinct groups and can be harnessed to identify specific gaps in treatment and care services at precise time points following diagnosis (1,3).

Existing evidence suggests youth living with HIV (YLHIV) experience worse HIV treatment outcomes at each stage in the HIV care continuum compared to adults (4–8). In sub-Saharan Africa, YLHIV are particularly vulnerable to suboptimal treatment outcomes (7,9). A 2016 meta-analysis conducted by Zanoni and colleagues found that just 14% of South African YLHIV aged 15 to 24 accessed antiretroviral treatment (ART) (10). Among those who accessed treatment, an estimated 83% were retained in care and 81% were virally suppressed, yielding an overall prevalence of suppression of 10% (10).

In line with the World Health Organization’s treatment recommendations, the South African government adopted a Universal Test and Treat (UTT) policy in September 2016 (11). This policy increased access to ART for all people living with HIV regardless of clinical stage and improved a number of HIV treatment outcomes among South African adults living with HIV (12). However, some studies suggest UTT increased attrition from care following treatment initiation (13). While little is known about the specific effects of this policy on treatment outcomes among YLHIV specifically, current data suggest poor retention in care and viral suppression persist in the population (14,15).

Addressing barriers to sustained engagement in care among YLHIV is critical if we are to end the HIV epidemic by 2030 (8,16). Yet, a limited number of studies have identified temporally precise opportunities for re-engaging this population following lapses in care (6). We draw on the longitudinal HIV care continuum framework to estimate the proportion of individuals in each stage of the HIV care continuum over time and the restricted mean time spent in each stage during the first year following diagnosis in a population of YLHIV in rural South Africa. We compare these outcomes by treatment era of diagnosis--pre- or post-UTT implementation--to illuminate the effects of this policy on longitudinal care trajectories and opportunities for re-engaging this priority population under current treatment recommendations.

## Methods

### Study site

This study was conducted in the Agincourt Health and Socio-Demographic Surveillance System study area (HDSS) in rural Mpumalanga Province, South Africa (17). This area is approximately 500 kilometers northeast of Johannesburg (17) and home to nearly 120,000 individuals (18). An estimated 27% of young women and 6% of young men ages 20 to 24 in this area are living with HIV (19). Access to public sector services and economic opportunities post-schooling is limited, contributing to a high degree of work-related migration, particularly among youth exiting the school system (20). Nine publicly funded health care facilities provide medical services to a majority of study area residents. Within these facilities, access to primary health care and ART is free of charge. However, patient wait times often exceed national standards and there is limited to no differentiated care for young people (21).

### Study population

We extracted data for all individuals aged 18 to 24 with a recorded HIV diagnosis in the HDSS-Clinic Link System (n=685), described in further detail below, between August 1^st^, 2015, and December 31^st^, 2018. Individuals who were not diagnosed in one of the nine publicly funded health care facilities used by residents of the Agincourt HDSS (n=251), had a viral load measurement below 400 copies/mL within seven days of diagnosis (n=29) or migrated into the Agincourt HDSS after their first HIV diagnosis (n=16) were excluded from the analysis to ensure participants were diagnosed and entered HIV care in an Agincourt HDSS facility.

### Data source

We used data from the Agincourt HDSS-Clinic Link System, previously described, to determine population clinical outcomes (22–25). Briefly, the Clinic Link System is a population-based clinical care database that covers consenting/assenting patients seeking HIV-specific services or chronic care in all nine publicly funded health care facilities used by study area residents. Data typists, stationed at each of the facilities since 2014, consent/assent patients seeking care on a daily basis. After obtaining written informed consent/assent, clinical visit data and patient demographic data are captured in the Clinic Link System and linked to corresponding records in the Agincourt HDSS census database. Data typists continually update clinical records data as individuals return for services.

Because the HDSS-Clinic Link System viral load data were occasionally missing, viral load measurements were supplemented using viral load data from the South African National Health Laboratory Service. The South African National Health Laboratory Service provides HIV diagnostic services to approximately 80% of South Africans and conducted more than 5 million viral load tests across 16 laboratories in 2018 (26,27).

Mortality and migration data were obtained from the Agincourt HDSS census database and linked to the HDSS-Clinic Link System data. The Agincourt HDSS database has been updated annually since 2000 and provides information on resident status and vital events such as migrations, births, and deaths (17).

### Measures

#### Linkage to care

Individuals were considered linked to care on the first of the following dates: results delivered for CD4 testing after HIV diagnosis, a follow-up visit with an indication of HIV treatment delivery, or a CD4 or viral load test after HIV diagnosis.

#### Loss to follow-up

Participants with no documented clinic visits for any given 90-day period following diagnosis were considered lost to follow-up (LTFU) on the first date the definition was met (i.e., the 90^th^ day following the most recent visit date). This definition is consistent with a lapse in medication coverage defined in the South African national HIV adherence guidelines (28). We also considered a 180-day clinic visit lapse definition of LTFU. Results from this analysis are presented in the Supplemental Material.

#### Treatment status

Participants were considered on ART the first date of any HIV treatment medication pickup. Individuals who had a suppressed viral load measurement prior to the first recorded ART pickup date (n=3) were considered on ART the same date as the suppressed viral load measurement.

#### Viral suppression status

Viral load measurements less than 400 copies/mL were considered virally suppressed (29,30).

#### Virologic failure

Viral load measurements of 1000 copies/mL and above subsequent to a suppressed viral load measurement were considered indicative of virologic failure (31,32).

#### Suboptimal treatment outcome

Because death (n=1) and virologic failure (n=3) were uncommon in the study population overall, we combined the competing events of LTFU, death, and virologic failure in a “suboptimal treatment outcomes” measure.

### Statistical analysis

To characterize the longitudinal HIV care continuum in the study population we utilized analytic methods similar to those formalized by Lesko et. al (3). These methods are designed to account for competing events and transitions into and out of multiple states over time (2). First, we fit a Fine and Grey sub-distribution proportional hazards models with no covariates and used the Breslow estimator to calculate the cumulative incidence of each of the following care continuum events following diagnosis: suboptimal treatment outcome prior to linkage to care; linkage to care; suboptimal treatment outcome prior to ART initiation (and after linkage to care); ART initiation; suboptimal treatment outcome prior to viral suppression (and after ART initiation); viral suppression; suboptimal treatment outcome following suppression (see S1 Table for additional details and competing events). Date of diagnosis served as the origin for each outcome and administrative censoring occurred on day 365 following diagnosis or February 1, 2019. Once an individual was LTFU, they were not permitted to reenter the study population.

We then estimated the proportion of the population in eight mutually exclusive stages of the HIV care continuum at each timepoint following diagnosis (i.e. on any day during the 365 days of follow-up):

1. Diagnosed with HIV- not yet linked to care
2. Suboptimal treatment outcome before linkage to care
3. Linked to care- ART naïve
4. Suboptimal treatment outcome after linkage to care but before ART initiation
5. On ART- virally non-suppressed
6. Suboptimal treatment outcome after ART initiation but before viral suppression
7. On ART- virally suppressed
8. Suboptimal treatment outcome after suppression

This was accomplished by adding and subtracting cumulative incidence curves (see S2 Table for equations). The proportion of individuals in each stage of the continuum is presented as a set of stacked curves that sum to one at each time point by design. The area between adjacent curves represents the restricted (as each participant is followed for a maximum of one year and all outcomes may not be observed) mean time spent in each stage over the one-year follow-up period. Given individuals who were LTFU were not permitted to reenter the study population, estimates of the mean time spent linked to care-ART naïve; on ART-virally non-suppressed; and on ART-virally suppressed, represent the mean time spent in each of these stages with no prior 90-or-more day gap in care. Potential care continuum transitions are specified in the S1 Figure.

Ultimately, we estimated 1) the proportion of participants in each stage of the continuum and 2) the restricted mean time spent in each stage of the continuum over the one-year period following diagnosis in the cohort overall and stratified by treatment era of diagnosis. We used inverse probability of treatment weights to account for meaningful differences in the distribution of sex and age at diagnosis between the two groups. We calculated differences in outcomes among those diagnosed pre- (referent) and post-UTT implementation and estimated the 95% Wald confidence intervals (CI) using the standard error of estimates obtained from 300 non-parametric resamples of the data (33). All analyses were conducted in SAS 9.4 (SAS Institute Inc., Cary, NC).

### Ethics

This study was approved by the University of North Carolina at Chapel Hill’s Institutional Review Board, the University of the Witwatersrand’s Human Research Ethics Committee, and the Mpumalanga Provincial Health Research Committee.

## Results

A total of 420 individuals were included. A majority were female (n=389; 92.6%) and diagnosed after the adoption of UTT (n=266; 63.3%) (Table 1). Median age at diagnosis was 22.2 (interquartile range [IQR]: 20.4-23.7) and median CD4 cell count at diagnosis or entry into care was 333 (IQR: 217-458) (Table 1).

**Table 1.** Characteristics of study participants, stratified by treatment era of diagnosis.

|  | **Total**<br>**(n=420)**<br>**n (%)** | **Pre-UTT**<br>**(n=154)**<br>**n (%)** | **UTT**<br>**(n=266)**<br>**n (%)** |
| --- | --- | --- | --- |
| Sex |  |  |  |
| Female | 389 (92.6) | 146 (94.8) | 243 (91.4) |
| Male | 31 (7.4) | 8 (5.2) | 23 (8.6) |
| Age at diagnosis (median/IQR) | 22.2 (20.4-23.7) | 22.3 (20.7-23.6) | 22.2 (20.3-23.6) |
| CD4 cell count at diagnosis/entry into care (median/IQR)<sup>†</sup> | 333 (217-458) | 329 (200-438) | 333.5 (220.5-482.5) |
| <200 cells/mm<sup>3</sup> | 74 (22.5) | 32 (24.8) | 42 (21.0) |
| ≥200 cells/mm<sup>3</sup> | 255 (77.5) | 97 (75.2) | 158 (79.0) |

Abbreviations: IQR- interquartile range; UTT- Universal Test and Treat
<sup>†</sup>missing baseline CD4 n=91

At day 90 post-diagnosis, 81.8% of all participants had ever linked to care, 61.6% had ever initiated ART, and 5.9% had ever achieved viral suppression (S2 Figure). Over one-third (33.8%) were LTFU on day 90 (S2 Figure). By 6 and 12 months post-diagnosis, 16.9% and 30.9% of all participants had ever achieved viral suppression, respectively (S2 Figure). By the end of the first year following diagnosis, 68.2% of all participants were LTFU, 0.2% had died, and 0.7% had experienced virologic failure. Participants spent a mean time of 19.5 days (95% CI: 16.0, 23.0) between diagnosis and linkage to care; 29.4 days (95% CI: 25.6, 33.3) linked to care but ART-naive; 107.4 days (95% CI: 95.9, 118.8) on ART but virally non-suppressed; and 53.7 days (95% CI: 45.2, 62.2) virally suppressed (Table 2).

**Table 2.**
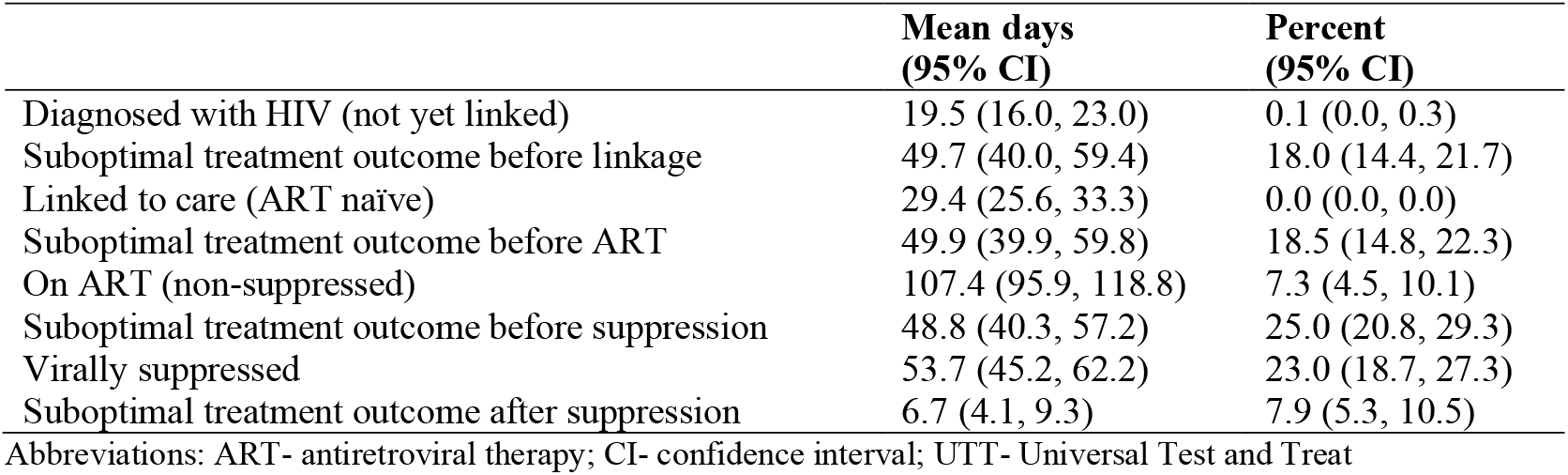
Time spent in each stage of care continuum over 1-year follow-up period and percent of individuals in each stage at end of follow-up.

|  | **Mean days**<br>**(95% CI)** | **Percent**<br>**(95% CI)** |
| --- | --- | --- |
| Diagnosed with HIV (not yet linked) | 19.5 (16.0, 23.0) | 0.1 (0.0, 0.3) |
| Suboptimal treatment outcome before linkage | 49.7 (40.0, 59.4) | 18.0 (14.4, 21.7) |
| Linked to care (ART naïve) | 29.4 (25.6, 33.3) | 0.0 (0.0, 0.0) |
| Suboptimal treatment outcome before ART | 49.9 (39.9, 59.8) | 18.5 (14.8, 22.3) |
| On ART (non-suppressed) | 107.4 (95.9, 118.8) | 7.3 (4.5, 10.1) |
| Suboptimal treatment outcome before suppression | 48.8 (40.3, 57.2) | 25.0 (20.8, 29.3) |
| Virally suppressed | 53.7 (45.2, 62.2) | 23.0 (18.7, 27.3) |
| Suboptimal treatment outcome after suppression | 6.7 (4.1, 9.3) | 7.9 (5.3, 10.5) |

Abbreviations: ART- antiretroviral therapy; CI- confidence interval; UTT- Universal Test and Treat

In weighted analyses, individuals diagnosed in the UTT era initiated ART more quickly after linkage to care (restricted mean time difference [MD] of time between linkage to care and ART initiation: -18.9; 95% CI: -27.3, -10.6), and spent more time over the one-year follow-up period on ART and virally suppressed when compared to those diagnosed in the pre-UTT era (MD of time spent on ART-virally non-suppressed: 12.7; 95% CI: -8.2, 33.7; MD of time spent virally suppressed: 17.7; 95% CI: 1.0, 34.4) (Table 3). Compared to their counterparts, time spent LTFU between diagnosis and linkage to care was greater among those diagnosed in the UTT era while time spent between linkage to care and ART initiation was greater among those diagnosed in the pre-UTT era (Table 3).

**Table 3.** Time spent in each stage of the HIV care continuum over 1-year follow-up period by treatment era of diagnosis.

|  | **Pre-UTT**<br>**Mean days**<br>**(95% CI)** | **UTT**<br>**Mean days**<br>**(95% CI)** | **Difference in mean**<br>**days**<br>**(95% CI)** |
| --- | --- | --- | --- |
| **Crude** |  |  |  |
| Diagnosed with HIV (not yet linked) | 17.3 (11.9, 22.7) | 21.4 (16.8, 26.0) | 4.1 (-2.9, 11.2) |
| Suboptimal treatment outcome before linkage | 40.8 (26.7, 55.0) | 54.7 (42.0, 67.3) | 13.8 (-5.0, 32.7) |
| Linked to care (ART naïve) | 41.3 (33.6, 49.0) | 22.3 (18.7, 26.0) | -18.9 (-27.3, -10.6) |
| Suboptimal treatment outcome before ART | 66.9 (47.9, 85.9) | 40.0 (29.2, 50.8) | -26.9 (-48.7, -5.1) |
| On ART (non-suppressed) | 99.2 (82.2, 116.3) | 111.8 (97.6, 125.9) | 12.5 (-8.5, 33.5) |
| Suboptimal treatment outcome before suppression | 51.3 (37.4, 65.2) | 47.2 (36.3, 58.1) | -4.1 (-21.5, 13.3) |
| Virally suppressed | 43.2 (31.0, 55.5) | 59.6 (47.9, 71.3) | 16.4 (-0.4, 33.1) |
| Suboptimal treatment outcome after suppression | 4.9 (1.4, 8.4) | 8.0 (4.4, 11.5) | 3.1 (-1.9, 8.0) |
| **Weighted<sup>†</sup>** |  |  |  |
| Diagnosed with HIV (not yet linked) | 16.9 (11.6, 22.1) | 21.3 (16.7, 25.8) | 4.4 (-2.5, 11.3) |
| Suboptimal treatment outcome before linkage | 39.8 (26.0, 53.6) | 54.0 (41.4, 66.7) | 14.2 (-4.3, 32.7) |
| Linked to care (ART naïve) | 41.5 (33.7, 49.3) | 22.2 (18.6, 25.9) | -19.3 (-27.7, -10.9) |
| Suboptimal treatment outcome before ART | 66.1 (47.2, 85.1) | 39.5 (28.8, 50.2) | -26.6 (-48.3, -5.0) |
| On ART (non-suppressed) | 99.4 (82.4, 116.4) | 112.1 (98.0, 126.2) | 12.7 (-8.2, 33.7) |
| Suboptimal treatment outcome before suppression | 54.0 (39.4, 68.6) | 47.7 (36.7, 58.7) | -6.3 (-24.4, 11.8) |
| Virally suppressed | 42.2 (30.1, 54.4) | 59.9 (48.2, 71.6) | 17.7 (1.0, 34.4) |
| Suboptimal treatment outcome after suppression | 5.0 (1.4, 8.7) | 8.2 (4.5, 11.9) | 3.2 (-2.0, 8.4) |

Abbreviations: ART- antiretroviral therapy; CI- confidence interval; UTT- Universal Test and Treat
<sup>†</sup>Weighted to account for differences in age at diagnosis and sex by treatment era of diagnosis

By the end of follow-up, 85.2% of participants diagnosed in the pre-UTT era had ever linked to care, 60.1% had ever initiated ART, and 26.0% had ever achieved viral suppression (Figure).

**Figure.**
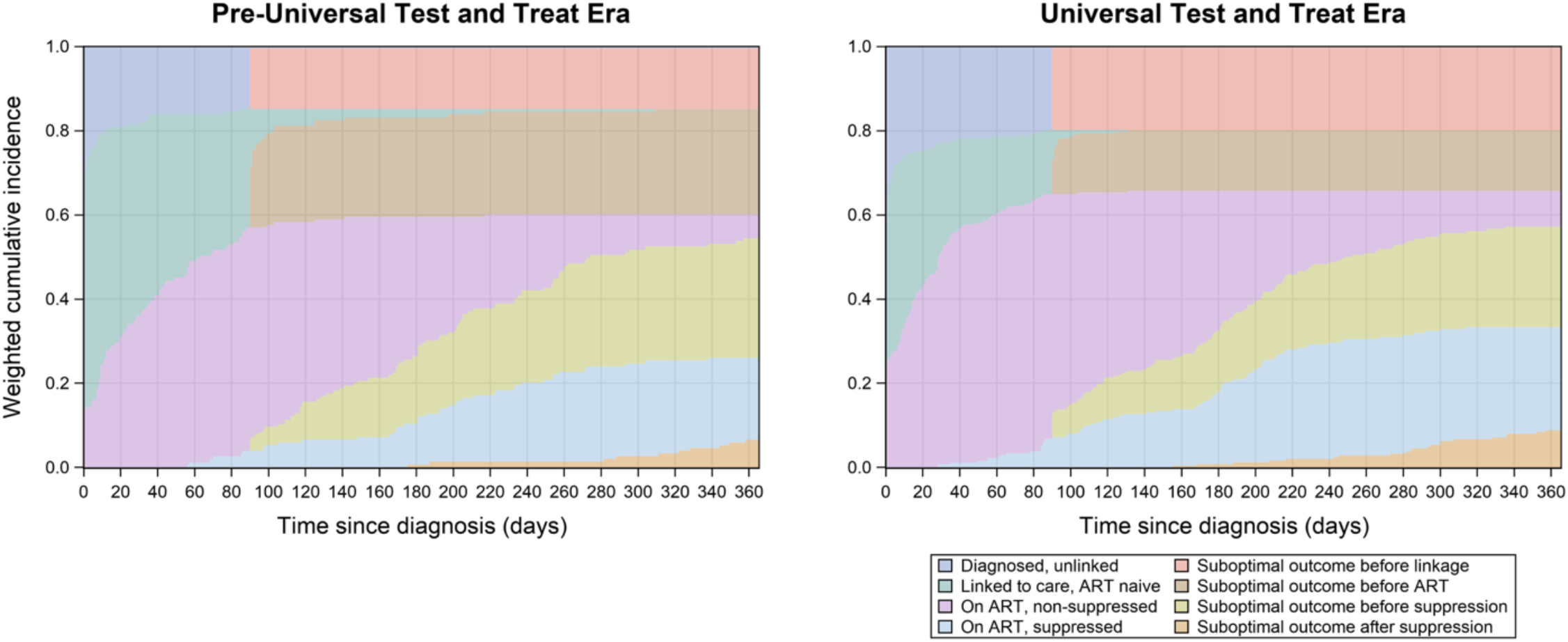
Cumulative incidence of HIV care outcomes over 1-year following diagnosis, stratified by treatment era. Abbreviations: ART- antiretroviral treatment; LTFU- lost to follow-up; UTT- Universal Test and Treat Weighted to account for differences in age at diagnosis and sex by era of diagnosis

Among those diagnosed in the UTT era, 80.2% had ever linked to care, 65.7% had ever initiated ART, and 33.3% had ever achieved viral suppression (Figure). On the last day of follow-up, 5.7% of participants diagnosed in the pre-UTT era were on ART but non-suppressed, and 19.5% were virally suppressed (Table 4; Figure). Among those diagnosed in the UTT era, 8.5% were on ART but non-suppressed, and 24.5% were virally suppressed (Table 4; Figure). Over 70% of participants diagnosed in the pre-UTT era, as compared to just over 60% in the post-UTT era experienced a suboptimal treatment outcome by the end of the first year following diagnosis with HIV (Table 4; Figure). Suboptimal outcomes between linkage to care and ART initiation were more common among those diagnosed in the pre-UTT versus UTT era by the end of study follow-up and sub-optimal outcomes between diagnosis and linkage to care were more common among those diagnosed in the UTT versus pre-UTT (Table 4; Figure). Approximately 25% of those diagnosed in both the pre-UTT and UTT eras were LTFU between ART initiation and viral suppression (Table 4; Figure).

**Table 4.**
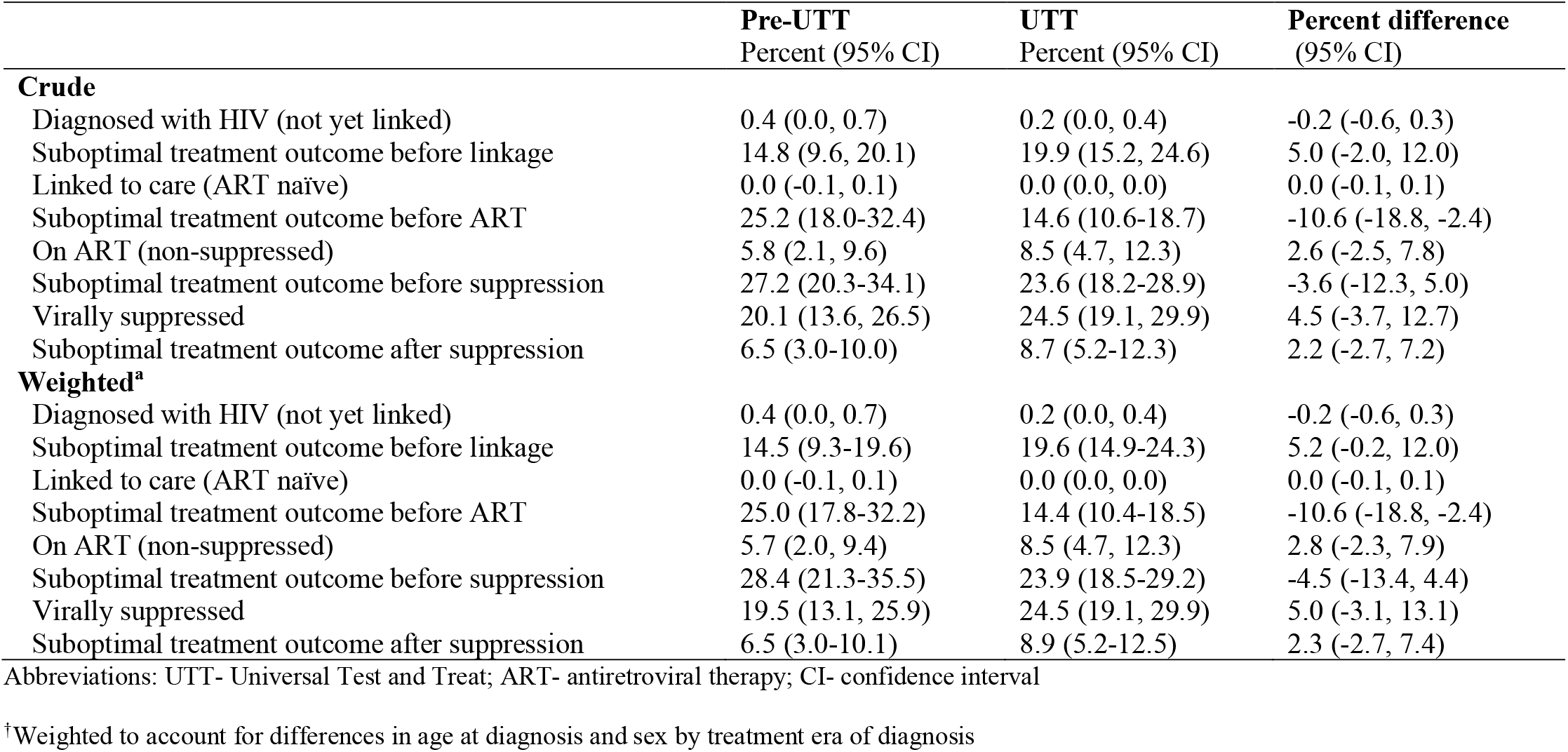
Proportion of participants in each stage of the HIV care continuum 1-year following diagnosis by treatment era of diagnosis.

|  | **Pre-UTT**<br>Percent (95% CI) | **UTT**<br>Percent (95% CI) | **Percent difference**<br>(95% CI) |
| --- | --- | --- | --- |
| **Crude** |  |  |  |
| Diagnosed with HIV (not yet linked) | 0.4 (0.0, 0.7) | 0.2 (0.0, 0.4) | -0.2 (-0.6, 0.3) |
| Suboptimal treatment outcome before linkage | 14.8 (9.6, 20.1) | 19.9 (15.2, 24.6) | 5.0 (-2.0, 12.0) |
| Linked to care (ART naïve) | 0.0 (-0.1, 0.1) | 0.0 (0.0, 0.0) | 0.0 (-0.1, 0.1) |
| Suboptimal treatment outcome before ART | 25.2 (18.0-32.4) | 14.6 (10.6-18.7) | -10.6 (-18.8, -2.4) |
| On ART (non-suppressed) | 5.8 (2.1, 9.6) | 8.5 (4.7, 12.3) | 2.6 (-2.5, 7.8) |
| Suboptimal treatment outcome before suppression | 27.2 (20.3-34.1) | 23.6 (18.2-28.9) | -3.6 (-12.3, 5.0) |
| Virally suppressed | 20.1 (13.6, 26.5) | 24.5 (19.1, 29.9) | 4.5 (-3.7, 12.7) |
| Suboptimal treatment outcome after suppression | 6.5 (3.0-10.0) | 8.7 (5.2-12.3) | 2.2 (-2.7, 7.2) |
| **Weighted<sup>a</sup>** |  |  |  |
| Diagnosed with HIV (not yet linked) | 0.4 (0.0, 0.7) | 0.2 (0.0, 0.4) | -0.2 (-0.6, 0.3) |
| Suboptimal treatment outcome before linkage | 14.5 (9.3-19.6) | 19.6 (14.9-24.3) | 5.2 (-0.2, 12.0) |
| Linked to care (ART naïve) | 0.0 (-0.1, 0.1) | 0.0 (0.0, 0.0) | 0.0 (-0.1, 0.1) |
| Suboptimal treatment outcome before ART | 25.0 (17.8-32.2) | 14.4 (10.4-18.5) | -10.6 (-18.8, -2.4) |
| On ART (non-suppressed) | 5.7 (2.0, 9.4) | 8.5 (4.7, 12.3) | 2.8 (-2.3, 7.9) |
| Suboptimal treatment outcome before suppression | 28.4 (21.3-35.5) | 23.9 (18.5-29.2) | -4.5 (-13.4, 4.4) |
| Virally suppressed | 19.5 (13.1, 25.9) | 24.5 (19.1, 29.9) | 5.0 (-3.1, 13.1) |
| Suboptimal treatment outcome after suppression | 6.5 (3.0-10.1) | 8.9 (5.2-12.5) | 2.3 (-2.7, 7.4) |

Abbreviations: UTT- Universal Test and Treat; ART- antiretroviral therapy; CI- confidence interval
<sup>a</sup>Weighted to account for differences in age at diagnosis and sex by treatment era of diagnosis

In analyses using the less restrictive LTFU definition, differences in the restricted mean time spent in each stage of the care continuum and the proportion in each stage at the end of the one-year follow-up period were attenuated (S3-5 Tables).

## Discussion

This study characterizes longitudinal HIV care continuum outcomes among YLHIV newly initiating HIV care in nine publicly funded health care facilities in rural South Africa pre- and post-implementation of the UTT policy. Overall, 82% of individuals had linked to care, 63% had initiated ART, and 31% had achieved viral suppression by one year following diagnosis. There were modest improvements in time spent on ART and virally suppressed among YLHIV diagnosed after the adoption of the UTT policy. However, the proportion of individuals diagnosed in the UTT era who had initiated ART, were retained in care, and were virally suppressed one year following diagnosis was well below the Joint United Nations’ Programme on HIV/AIDS’ 95-95-95 goal. A substantial proportion of individuals diagnosed in the UTT era disengaged from care in the periods between diagnosis and linkage to care and ART initiation and viral suppression.

Approximately 66% of individuals diagnosed during the UTT era initiated ART in the first year following diagnosis. The mean time spent suppressed during this period was 60 days and just 25% of individuals were suppressed at the end of follow-up. This represents a substantial gap in sustained retention on ART and viral suppression among YLHIV diagnosed in the era of UTT that must be urgently addressed.

Implementation of UTT appeared to influence the timing of care disengagement among YLHIV in this study. Specifically, those diagnosed in the pre-UTT era were more likely to be LTFU between linkage to care and ART initiation while those diagnosed in the UTT era were more likely to be LTFU between diagnosis and linkage to care. This suggests individuals diagnosed during the UTT era may have been less likely to return to care to start ART if they were not initiated the same day they were diagnosed. Existing evidence suggests low ART readiness may impede retention in care following diagnosis in the UTT era (34). For example, a study of adults immediately referred for ART in South Africa found that individuals who did not expect to receive a positive HIV diagnosis had significantly lower odds of ART readiness than others (adjusted odds ratio 0.26; 95% CI: 0.09, 0.78) and that the odds of linkage to care among those expressing treatment readiness were 2.97 times that in individuals who were not ready to initiate ART (34,35).

These findings have important implications for HIV policy and programming for South African YLHIV in the UTT era. First, improved counseling and linkage to care at the point of diagnosis with HIV may be critical to the success of same-day ART initiation policies. Second, increased resources in the UTT era should be spent on retention in care efforts for YLHIV, particularly in the periods between diagnosis and linkage to care and ART initiation and viral suppression. Longitudinal counseling following diagnosis has been shown to improve linkage to care in other settings across sub-Saharan Africa (36,37). For example, in a cluster randomized controlled trial exploring the effects of a counseling intervention on treatment outcomes among people living with HIV in Uganda, those in the intervention arm were significantly more likely to link to care compared to those in the control arm (37). Differentiated, youth-friendly medical care and programs that target improvements in young people’s self-esteem, social support, and overall psychosocial well-being may also be important to improving care outcomes for YLHIV (38–44). A retrospective cohort study of youth initiating ART at 37 facilities in South Africa demonstrated just 19% of study participants receiving support services were non-suppressed 5-years after ART initiation compared to 37% of those who did not receive community based-services (44). Care models such as these should be prioritized for South African YLHIV particularly in the critical first year following diagnosis.

Increased attention must also center on monitoring care services for this group, specifically. While the cross-sectional HIV care cascade (45) is useful in describing the proportion of individuals in each stage of the care continuum at a specific point in time, it provides a mere snapshot of the true patient experience (2,46). Longitudinal HIV care cascades allow for a nuanced exploration of population-level outcomes over time (1,2) and can aid in assessing the effectiveness of HIV care programs as individuals progress through each stage of the continuum. Ultimately, these cascades can be harnessed to identify specific gaps in treatment and care services at precise time points following diagnosis (1,3).

This analysis had limitations. First, viral load monitoring is recommended just once per year under the South African National HIV Treatment guidelines (28). Because we were primarily interested in assessing HIV treatment outcomes in the first year following an individual’s diagnosis, most participants had one opportunity to achieve viral suppression during the study period. Among those who achieved viral suppression, true time to viral suppression may have been shorter but went uncaptured because of infrequent viral load monitoring. Similarly, virologic failure among individuals who achieved suppression may have been under-captured due to infrequent viral load monitoring. Misclassified person-time could subsequently result in biased effect estimates. Second, mortality and migration data were only accessible from 2014-2017. Individuals who died in 2018 and were not yet administratively censored or LTFU before the time of death would have misclassified person-time. Given just one participant died in the 2014-2017 follow-up period, this was a minor concern. Lastly, the nature of the Clinic Link System prevented us from reliably ascertaining the reason for individuals’ LTFU. It is possible that individuals who were classified as LTFU transferred into care at a clinic outside the nine included health care facilities, as documented in other studies (24,25). Individuals who transferred care and were classified as LTFU may have progressed through additional stages in the HIV care continuum during the one-year follow-up period, though we do not expect differential migration with respect to treatment era.

## Conclusions

Implementation of the World Health Organization’s UTT policy yielded modest improvements in the time spent on ART and virally suppressed among South African YLHIV. However, with just 66% of YLHIV diagnosed in the UTT era initiating ART, and just 25% virally suppressed one year following diagnosis, meeting UNAIDS 95-95-95 targets remains a challenge. HIV treatment programs and policies for YLHIV in the UTT era should specifically center on improving longitudinal care outcome monitoring and retention in care in the periods immediately following diagnosis and ART initiation.

## Data Availability

The datasets analyzed during the current study are not publicly available due to the sensitive nature of HIV treatment and care data. Data may be made available from the study team upon reasonable request.

## Abbreviations

ART: anti-retroviral therapy
HDSS: Health and Socio-Demographic Surveillance System study area
HIV: human-immunodeficiency virus
LTFU: lost-to follow-up
UNAIDS: United Nations’ Joint Programme on HIV/AIDS
UTT: Universal Test and Treat policy
YLHIV: youth living with HIV

## Declarations

### Ethics approval and consent to participate

This study was approved by the University of North Carolina at Chapel Hill’s Institutional Review Board, the University of the Witwatersrand’s Human Research Ethics Committee, and the Mpumalanga Provincial Health Research Committee. All individuals provided written informed consent or assent for inclusion of their electronic health records in the Agincourt-HDSS Clinic Link System.

### Consent for publication

Not applicable.

### Availability of data and materials

The datasets analyzed during the current study are not publicly available due to the sensitive nature of HIV treatment and care data. Data may be made available from the study team on reasonable request.

### Authors contributions

LMF- Conceptualization; Funding; Project administration; Data curation; Formal analysis; Methodology; Writing- original draft; JKE- Conceptualization; Methodology; Writing- revising & editing; NM-Project administration; Data curation; Writing- revising & editing; FXGO- Project administration; Methodology; Writing- revising & editing; NH- Funding; Writing- revising & editing; BP- Methodology; Writing- revising & editing; AM- Methodology; Writing- revising & editing; KM- Methodology; Writing- revising & editing; CWK- Data curation; Project administration; Writing- revising & editing; MKD- Data curation; Writing- revising & editing; SAL- Funding; Methodology; Writing- revising & editing; KK- Funding; Methodology; Project administration; Writing- revising & editing; AP- Conceptualization; Funding; Methodology; Writing- revising & editing.

## Acknowledgments

The authors would like to thank all the individuals who participated in this study.

## Additional Files

S1 Table: Events of interest in longitudinal HIV care continuum

S2 Table: Equations for estimating the proportion of individuals in each stage of the continuum at any given time point during study follow-up

S1 Figure: Framework for flow through the longitudinal HIV care continuum

S2 Figure. Cumulative incidence of HIV care outcomes over the 1-year follow-up period in study population overall

S3 Table. Time spent in each stage of care continuum over 1-year follow-up period and percent of individuals in each stage at end of follow-up^a^

S4 Table. Time spent in each stage of the HIV care continuum over 1-year follow-up period by treatment era of diagnosis^a^

S5 Table. Proportion of participants in each stage of the HIV care continuum 1-year following diagnosis by treatment era of diagnosis^a^

