## Supplementary material for "Understanding the effects of Universal Test and Treat on longitudinal HIV care continuum outcomes among South African youth: a retrospective cohort study": S1 Table

**Supplemental Digital Content**

##### Supplemental Table 1. Events of interest in longitudinal HIV care continuum

| **Outcome** | **Definition** | **Competing event** |
| --- | --- | --- |
| Loss to follow-up (LTFU) or death before linkage to care (R_1_) | Death date (prior to care initiation) | Linkage to care |
|  | No HIV clinic visit dates for any 90-day interval (before care) |  |
| Linkage to care (R_2_) | First visit when HIV-related medication or ART was diagnosed, or VL or CD4 count test was conducted | LTFU or death before linkage to care |
| Loss to follow-up or death before ART initiation (after linkage to care) (R_3_) | Death date (prior to ART initiation)  No HIV clinic visit dates for any 90-day interval (before ART) | LTFU or death before linkage to care; ART initiation |
| ART initiation (R_4_) | First date of any HIV treatment medication pick up on, or following, their date of HIV diagnosis | LTFU or death before linkage to care or ART initiation |
| Loss to follow-up or death before suppression (after ART initiation) (R_5_) | Death date (prior to suppression)  No HIV clinic visit dates for any 90-day interval (prior to suppression) | LTFU or death before linkage to care or ART initiation; viral suppression |
| Viral suppression (R_6_) | Viral load <400 copies/mL | LTFU or death before linkage care, ART initiation, or viral suppression |
| Loss to follow-up, death, or virologic failure after suppression (R_7_) | Death date (prior to virologic failure),  No HIV clinic visit dates for any 90-day interval (after suppression), or viral load $\geq$400 copies/mL subsequent to viral suppression | LTFU or death before linkage to care, ART initiation, or viral suppression |

ART- antiretroviral therapy; LTFU- lost to follow-up
