## Supplementary material for "Understanding the effects of Universal Test and Treat on longitudinal HIV care continuum outcomes among South African youth: a retrospective cohort study": S2 Table

**Supplemental Digital Content**

##### Supplemental Table 2. Equations for estimating the proportion of individuals in each stage of the continuum at any given time point during study follow-up

Diagnosed with HIV, not yet in care	P(D)	=	1-R_1_(x)-R_2_(x)

Suboptimal outcome before linkage to care	P(L_1_)	=	R_1_(x)

In care, ART naïve	P(C)	=	R_2_(x)-R_3_(x)-R_4_(x)

Suboptimal outcome before ART initiation	P(L_2_)	=	R_3_(x)

On ART, virally non-suppressed	P(A)	=	R_4_(x)- R_5_(x)-R_6_(x)

Suboptimal outcome before suppression	P(L_3_)	=	R_5_(x)

On ART, virally suppressed	P(S)	=	R_6_(x)-R_7_(x)

Suboptimal outcome after suppression	P(L_4_)	=	R_7_(x)

ART- antiretroviral treatment
