## Supplementary material for "Understanding the effects of Universal Test and Treat on longitudinal HIV care continuum outcomes among South African youth: a retrospective cohort study": S1 Figure

**Supplemental Digital Content**

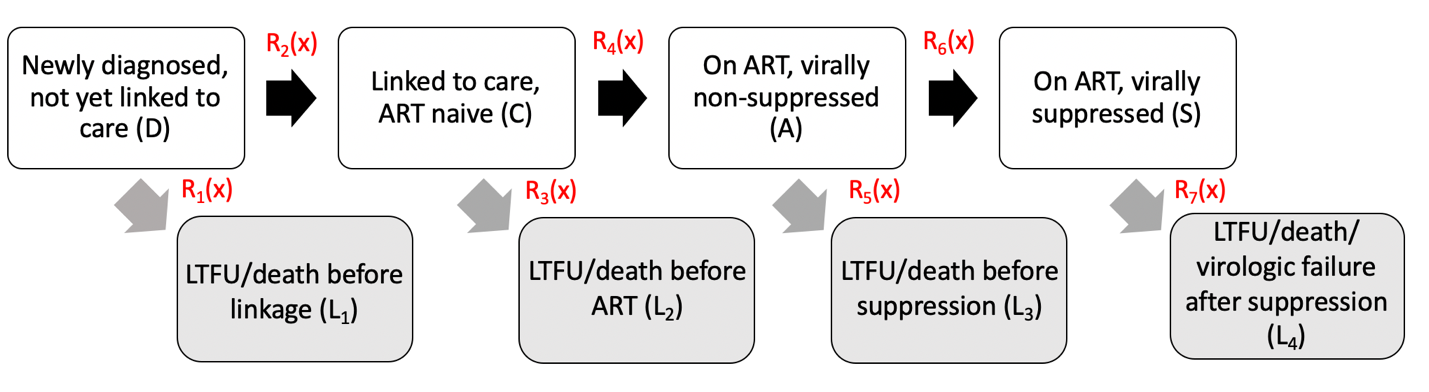

**Supplemental Figure 1. Framework for flow through the longitudinal HIV care continuum**

ART-antiretroviral treatment initiation; LTFU- loss to follow-up
