## Supplementary material for "Understanding the effects of Universal Test and Treat on longitudinal HIV care continuum outcomes among South African youth: a retrospective cohort study": S2 Figure

**Supplemental Digital Content**

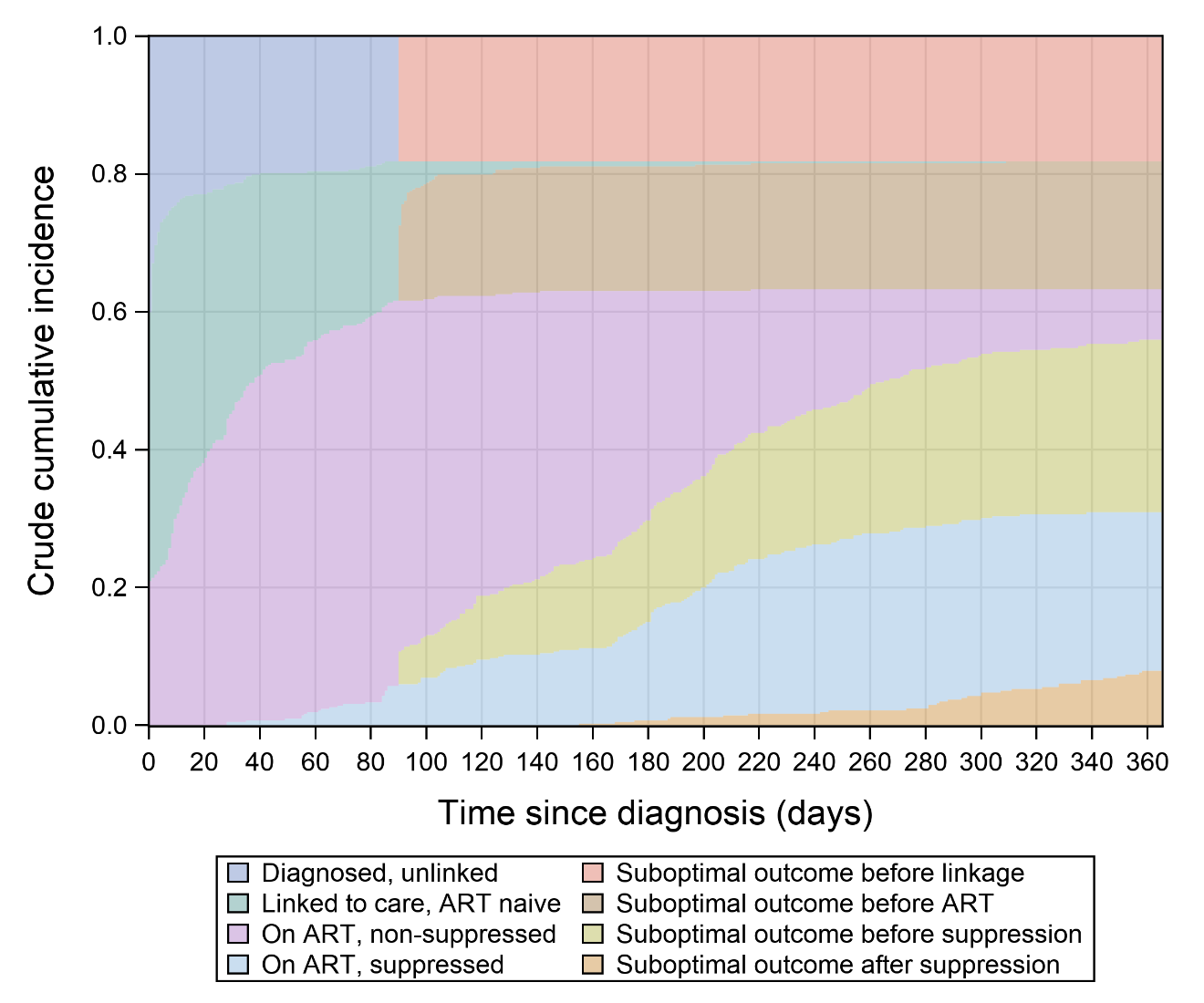

**Supplemental Figure 2. Cumulative incidence of HIV care outcomes over the 1-year follow-up period in study population overall**

ART- antiretroviral treatment; LTFU- lost to follow-up; UTT- Universal Test and Treat
