## Supplementary material for "Understanding the effects of Universal Test and Treat on longitudinal HIV care continuum outcomes among South African youth: a retrospective cohort study": S3 Table

**Supplemental Digital Content**

**Supplemental Table 3. Time spent in each stage of care continuum over 1-year follow-up period and percent of individuals in each stage at end of follow-up^a^**

|  | **Mean days**  **(95% CI)** | **Percent (95% CI)** |
| --- | --- | --- |
| Diagnosed with HIV, not yet linked | 35.3 (28.9, 41.8) | 0.1 (0.0, 0.3) |
| Suboptimal treatment outcome before linkage | 31.2 (25.3, 37.1) | 16.9 (13.5, 20.2) |
| Linked to care, ART naïve | 44.2 (37.6, 50.9) | 0.2 (-0.2, 0.7) |
| Suboptimal treatment outcome before ART | 23.2 (17.4, 28.9) | 13.3 (9.9, 16.7) |
| On ART, non-suppressed | 142.0 (130.3, 153.8) | 15.6 (11.9, 19.3) |
| Suboptimal treatment outcome before suppression | 18.9 (14.4, 23.3) | 15.9 (12.2, 19.6) |
| On ART, suppressed | 67.9 (58.4, 77.4) | 35.4 (30.5, 40.4) |
| Suboptimal treatment outcome after suppression | 2.3 (1.0, 3.7) | 2.5 (1.0, 4.0) |

ART- antiretroviral therapy; CI- confidence interval; UTT- Universal Test and Treat

^a^Loss to follow up defined as no documented clinic visits for any given 180-day period following diagnosis
