## Supplementary material for "Understanding the effects of Universal Test and Treat on longitudinal HIV care continuum outcomes among South African youth: a retrospective cohort study": S4 Table

**Supplemental Digital Content**

**Supplemental Table 4. Time spent in each stage of the HIV care continuum over 1-year follow-up period by treatment era of diagnosis^a^**

|  | **Pre-UTT**  **Mean days**  **(95% CI)** | **UTT**  **Mean days**  **(95% CI)** | **Difference in mean days**  **(95% CI)** |
| --- | --- | --- | --- |
| **Crude** |  |  |  |
| Diagnosed with HIV, not yet linked | 30.7 (20.3, 41.1) | 38.6 (30.3, 47.0) | 8.0 (-5.6, 21.5) |
| Suboptimal treatment outcome before linkage | 27.4 (17.9, 37.0) | 33.3 (25.6, 41.0) | 5.8 (-6.8, 18.4) |
| Linked to care, ART naïve | 57.7 (44.9, 70.4) | 36.1 (29.0, 43.2) | -21.6, -35.9, -7.2) |
| Suboptimal treatment outcome before ART | 24.6 (15.3, 34.0) | 22.4 (15.4, 29.4) | -2.2 (-13.6, 9.2) |
| On ART, non-suppressed | 138.6 (119.6, 157.6) | 143.5 (128.9, 158.1) | 4.9 (-18.6, 28.4) |
| Suboptimal treatment outcome before suppression | 21.4 (14.5, 28.3) | 17.6 (12.3, 22.8) | -3.8 (-12.2, 4.5) |
| On ART, suppressed | 62.8 (48.5, 77.2) | 70.8 (59.1, 82.5) | 8.0 (-9.8, 25.7) |
| Suboptimal treatment outcome after suppression | 1.7 (0.2, 3.3) | 2.7 (0.7, 4.7) | 1.0 (-1.7, 3.6) |
| **Weighted^b^** |  |  |  |
| Diagnosed with HIV, not yet linked | 29.9 (19.7, 40.0) | 38.3 (30.0, 46.6) | 8.4 (-4.9, 21.7) |
| Suboptimal treatment outcome before linkage | 26.8 (17.5, 36.0) | 32.9 (25.2, 40.5) | 6.1 (-6.2, 18.5) |
| Linked to care, ART naïve | 57.9 (45.0, 70.9) | 35.8 (28.7, 42.9) | -22.2 (-36.7, -7.6) |
| Suboptimal treatment outcome before ART | 24.7 (15.3, 34.0) | 22.1 (15.2, 29.1) | -2.5 (-13.9, 8.8) |
| On ART, non-suppressed | 139.7 (120.7, 158.8) | 144.2 (129.6, 158.9) | 4.5 (-19.2, 28.2) |
| Suboptimal treatment outcome before suppression | 22.2 (15.1, 29.3) | 17.8 (12.4, 23.1) | -4.4 (-13.1, 4.2) |
| On ART, suppressed | 62.0 (47.6, 76.3) | 71.1 (59.3, 82.9) | 9.2 (-8.5, 26.9) |
| Suboptimal treatment outcome after suppression | 1.8 (0.2, 3.5) | 2.8 (0.7, 5.0) | 1.0 (-1.8, 3.8) |

Abbreviations: UTT- Universal Test and Treat; ART- antiretroviral therapy; CI- confidence interval

^a^Loss to follow up defined as no documented clinic visits for any given 180-day period following diagnosis

^b^Weighted to account for differences in age at diagnosis and sex by treatment era of diagnosis
