## Supplementary material for "Understanding the effects of Universal Test and Treat on longitudinal HIV care continuum outcomes among South African youth: a retrospective cohort study": S5 Table

**Supplemental Digital Content**

**Supplemental Table 5. Proportion of participants in each stage of the HIV care continuum 1-year following diagnosis by treatment era of diagnosis^a^**

|  | **Pre-UTT**  Percent (95% CI) | **UTT**  Percent (95% CI) | **Percent difference**  (95% CI) |
| --- | --- | --- | --- |
| **Crude** |  |  |  |
| Diagnosed with HIV, not yet linked | 0.4 (0.0, 0.8) | 0.2 (0.0, 0.5) | -0.2 (-0.6, 0.3) |
| Suboptimal treatment outcome before linkage | 14.8 (9.3, 20.4) | 18.0 (13.7, 22.2) | 3.1 (-3.9, 10.2) |
| Linked to care, not on ART | 0.6 (-0.6, 1.9) | 0.0 (0.0, 0.0) | -0.6 (-1.9, 0.6) |
| Suboptimal treatment outcome before ART | 14.2 (8.4, 20.0) | 12.8 (8.6, 16.9) | -1.5 (-8.5, 5.5) |
| On ART, non-suppressed | 14.2 (8.8, 19.6) | 16.6 (11.9, 21.2) | 2.3 (-4.4, 9.0) |
| Suboptimal treatment outcome before suppression | 19.4 (13.1, 25.7) | 13.5 (9.3, 17.7) | -5.9 (-13.4, 1.5) |
| On ART, suppressed | 33.7 (25.9, 41.4) | 36.6 (30.5, 42.6) | 2.6 (0.1, 5.1) |
| Suboptimal treatment outcome after suppression | 2.6 (0.0, 5.1) | 2.4 (0.6, 4.3) | -0.1 (-3.3, 3.1) |
| **Weighted** |  |  |  |
| Diagnosed with HIV, not yet in care | 0.3 (0.0, 0.7) | 0.2 (0.0, 0.5) | -0.1 (-0.5, 0.3) |
| Suboptimal treatment outcome before linkage | 14.5 (9.1, 19.9) | 17.8 (13.5, 22.0) | 3.3 (-3.6, 10.2) |
| Linked to care, not on ART | 0.6 (-0.6, 1.9) | 0.0 (0.0, 0.0) | -0.6 (-1.9, 0.6) |
| Suboptimal treatment outcome before ART | 14.4 (8.5, 20.2) | 12.6 (8.5, 16.6) | -1.8 (-8.9, 5.2) |
| On ART, non-suppressed | 14.1 (8.7, 19.5) | 16.7 (12.0, 21.3) | 2.6 (-4.1, 9.2) |
| Suboptimal treatment outcome before suppression | 20.3 (13.7, 26.9) | 13.7 (9.4, 17.9) | -6.6 (-14.3, 1.1) |
| On ART, suppressed | 33.1 (25.3, 40.9) | 36.5 (30.4, 42.6) | 3.4 (-6.2, 13.1) |
| Suboptimal treatment outcome after suppression | 2.7 (0.0, 5.3) | 2.5 (0.6, 4.5) | -0.1, (-3.4, 3.2) |

ART- antiretroviral therapy; CI- confidence interval; UTT- Universal Test and Treat

^a^Loss to follow up defined as no documented clinic visits for any given 180-day period following diagnosis

^b^Weighted to account for differences in age at diagnosis and sex by treatment era of diagnosis
